# Associations of endogenous and exogenous hormonal exposures and cardiovascular disease in women – A FinnGen study

**DOI:** 10.64898/2026.03.13.26348304

**Authors:** Tia-Marje Korhonen, Takiy-Eddine Berrandou, Laura Joensuu, FinnGen, Jari Laukkanen, Elina Sillanpää, Nabila Bouatia-Naji, Eija K Laakkonen

**Affiliations:** Faculty of Sport and Health Sciences, Gerontology Research Center, University of Jyväskylä, Jyväskylä, FINLAND; Université Paris Cité, Paris Cardiovascular Research Center, Inserm, Paris, FRANCE; Wellbeing Services County of Central Finland, Jyväskylä, FINLAND; Institute of Clinical Medicine, University of Eastern Finland, Kuopio, FINLAND

**Keywords:** menopause, menarche, reproductive span, estrogen exposure, menopausal hormone therapy, hormonal contraceptives, stroke, hypertension, coronary heart disease

## Abstract

**Background:** Earlier age at menopause and shorter reproductive span (time from menarche to menopause) have been linked to an increased risk of cardiovascular diseases (CVD), presumably because of limited lifetime exposure to endogenous estrogen. Our intention is to determine whether genetic liability to earlier vs later menarche and menopause are associated with risk of common cardiovascular diseases in women. We also aim to investigate effects of exogenous estrogen exposure in the form of systemic hormonal contraceptive and menopausal hormone therapy use.

**Methods and results:** We determined GWAS summary statistics for age at menopause, age at menarche, and length of the reproductive period with data derived from UK Biobank (UKBB) European-ancestry participants. We calculated polygenic scores for women from FinnGen study (N=184 132) that indirectly capture genetic variation in endogenous estrogen exposure: age at menopause, age at menarche, and length of reproductive span. We also investigated exogenous hormone exposure associated with use of menopausal hormone therapy or systemic hormonal contraceptives. We used Cox proportional hazards model to investigate associations between PGSes and exogenous estrogen exposure and risk of hypertension, stroke, and coronary heart disease (CHD) events. Median follow-up time was 25.8 years. During the follow-up 56 143 women experienced hypertension, 18 200 women experienced strokes, and 13 879 women experienced major CHD events. Genetic liability to later menopause and longer reproductive span were weakly associated with a higher risk of stroke: in models adjusted with smoking and BMI, the hazard ratio (HR) per one standard deviation increase in PGS was 1.03 [1.01–1.05] for both. Menopausal hormone therapy use was associated with lower risk of stroke (HR 0.85 [0.82 – 0.89]) and CHD (HR 0.80 [0.76 – 0.84]). Systemic hormonal contraceptive use was associated with lower risk of hypertension (HR 0.96 [0.93 – 0.99]), stroke (HR 0.84 [0.80 – 0.99]) and CHD events (HR 0.83 [0.78–0.89]).

**Conclusions:** Although observational evidence consistently associates a longer reproductive span with lower cardiometabolic risk, the polygenic component of metabolic timing (PGSes for reproductive span and age at menopause) showed the opposite direction of association. This discrepancy likely reflects the fact that these scores capture genetic pathways only partially overlapping with phenotypic lifetime estrogen exposure. Importantly, the observed cardioprotective associations of menopausal hormone therapy and systemic hormonal contraception underscore that genetic predisposition and exogenous hormonal exposure represent distinct biological dimensions relevant to cardiovascular risk in women.

**Clinical Perspective:** *What Is New?:*

- We combined genetic data and information of register-based hormone purchases from over 180 000 women to investigate associations between endogenous and exogenous estrogen exposure and cardiovascular diseases.

*What Are the Clinical Implications?:*

- Our results suggest that genetic predisposition to a later natural menopause and longer reproductive span are not protective towards cardiovascular diseases.
- Exogenous hormones were associated with lower long-term risk of stroke and coronary heart disease events. This implies that even if the current use of exogenous estrogen may increase stroke risk, the long-term stroke risk may decrease compared to women who never used hormones.

## Introduction

There are major sex differences in several cardiovascular disease (CVD) traits. Women have specific forms and aetiology of cardiovascular diseases, including female-typical heart failure subtypes and cardiomyopathies [1]. Female sex hormones have a large effect on women’s cardiovascular risks [2]. Women have a higher lifetime risk of stroke compared to men [3] and worse outcomes [4]. Hypertension is the most prevalent risk factor for stroke [5]. Women have slightly lower blood pressure than men of the same age [6], but women’s risk for hypertension and stroke begins to increase around the menopause transition [5,6]. Estrogen modulates blood pressure and lipid levels and is considered to be cardioprotective [7]. The menopausal decline in estrogen levels is associated with adverse changes in clinical cardiovascular biomarkers [8, 9], blood metabolites [10], body composition [11,12], and vascular health [13,14], which may contribute to an increased risk of CVDs.

Cardiovascular risk factors in women are associated with the length of estrogen exposure over the lifespan. Early menopause, occurring before age 45, increases the risk of hypertension and stroke [5,15]. It has also been linked to an increased risk of CHD and CVD mortality and all-cause mortality [15,16]. Even menopause at the younger end of the normal range – that is, between 45 and 49 years – has been associated with a higher likelihood of first cardiovascular event before the age of 60 years [17]. Consistent with this gradient, each 1-year decrease in age at menopause has been shown to be associated with 2% higher risk of incident CDH [18]. Menarche typically occurs between ages 12 to 14. Typical reproductive span, i.e. time from menarche to menopause, is 33 to 39 years. Stroke risk is higher among women who have early (≤10 years) or late (≥16 years) menarche or have a reproductive life span of less than 30 years [5]. In a population study [19] of reproductive span and cardiovascular outcomes, a shorter reproductive period (lowest tertile) higher cardiovascular event risk while a longer reproductive period (highest tertile) was protective. Early menarche has been associated with higher BMI and higher risk of hypertension and CVDs [20, 21].

A substantial number of women use exogenous hormones at some point in their lives in the form of oral contraceptives or hormone replacement therapy. Long-term consequences and potential risks of exogenous hormones on women’s health require further study. Prior research shows an increased risk of stroke with oral contraceptive pill use [22, 23]. Menopausal hormone therapy (MHT) use has been associated with increased risk of ischemic heart disease, stroke, and venous thromboembolism [24, 25, 26], but the effects are strongly dependent on the formulation and delivery method [27]. In a recent study, tibolone and oral combined continuous therapy were associated with ischemic heart disease, while transdermal oestrogen did not show increased risks of CVDs [24].

In the current study, the main objective was to investigate associations between endogenous and exogenous estrogen exposure and CVDs prevalent in women, including hypertension, stroke, and CHD. We conducted survival analyses using Cox proportional hazard models in the FinnGen database, which includes close to 10% of Finland’s population. In the absence of phenotypic data on menarche, menopause, and reproductive span, we calculated polygenic scores (PGS) for age at menarche, age at menopause, and length of reproductive span and used them as proxies for these traits to test how they associate with hypertension, stroke, and major CHD events among Finnish population. To account for exogenous hormone exposure, we investigated how information on systemic hormonal contraceptive and MHT use, originating from the Finnish high-quality medical register data, was associated with the risk of CVDs.

## Methods

### Study design and subjects

The FinnGen study is a large-scale genomics initiative that has analyzed over 500,000 Finnish biobank samples (close to 10% of Finnish population) and correlated genetic variation with health data to understand disease mechanisms and predispositions. FinnGen combines genomic information with high-quality population health registries (www.finngen.fi/fi). FinnGen is enriched with disease cases from hospital biobanks and disease specific legacy cohorts deposited to Finnish biobanks.

Polygenic scores were computed for individuals in FinnGen R12 database (500 348 participants, 282 064 women). To focus on diseases that are related to long-term hormonal exposures, we started with women who were 40 years or older. Since we wanted to investigate natural menopause, we excluded women who had both ovaries surgically removed. We further excluded women who had any cardiovascular disease before age 40. This left us with 184 132 women for the analysis.

### Outcomes

As cardiovascular outcomes we used predefined endpoints from Finngen R12 database [https://www.finngen.fi/en/researchers/clinical-endpoints] including hypertension (I9_HYPTENS), stroke (C_STROKE), and major CHD event (I9_CHD). Definitions of the FinnGen endpoints used are included in Supplemental Table 1. During the median follow-up period of 25.8 years and total follow up time of over 4.7 million person years, 56 143 women experienced hypertension, 18 200 women experienced strokes, and 13 879 women experienced major CHD events.

### Endogenous hormonal exposure

To obtain genetic proxies of lifelong endogenous hormonal timing in FinnGen, we constructed PGS for age at menarche, age at menopause, and length of the reproductive period (age at menopause minus age at menarche) and tested their associations with the study outcomes. PGS were trained using GWAS performed in UK Biobank women of European genetic ancestry using age at menarche (data-field 2714) and age at menopause (data-field 3581, age at last menstrual period). After excluding non-informative responses and implausible values (menarche 10–18 years; menopause 40–60 years) and requiring both measures, 109,758 women were retained for each trait. GWAS were run with PLINK2 under an additive linear regression model adjusted for the first 10 genetic principal components, restricting to well-imputed common variants (MAF ≥ 0.01; imputation INFO > 0.8).

Using these GWAS summary statistics, we trained PGS with LDAK (v6.1) MegaPRS under a BayesR prior [28]. We first computed SNP–SNP correlations in a 10,000-sample UK Biobank European-ancestry LD reference panel using LDAK --calc-cors, restricting to autosomal HapMap3 variants and excluding variants with MAF < 0.01 in the LD reference and region with high LD; correlations were computed within 4 Mb windows. MegaPRS was then run on the resulting correlation files using BayesR model with LDAK’s default shrinkage and pseudo cross-validation settings to select the best hyperparameters. The training set comprised 1,248,174 HapMap3 variants available in the LD reference, of which 1,247,934 had corresponding GWAS summary statistics and were used to estimate final SNP weights. The resulting SNP weights were applied to FinnGen participants using PLINK 2 (--score) [29], with alleles aligned to the GWAS effect allele. For any missing genotype at a scored variant, PLINK2 substituted the variant’s mean genotype dosage computed in the scoring dataset (default behavior). Scores were then standardized within the analysis sample.

### Exogenous hormonal exposure

Drug use was derived from drug purchase and reimbursement registers of the Social Insurance Institution of Finland Kela, a system that records all reimbursable drug purchases made at Finnish pharmacies. Rather than looking at specific drugs, hormone use was determined from the anatomical therapeutic chemical (ATC) classifications of drugs used by the study participants. The ATC classification is a system used to categorize drugs and active substances based on their anatomical target, therapeutic use, and chemical properties. Women were considered systemic hormonal contraceptive users if they had used drugs in ATC classification G03A “Hormonal contraceptives for systemic use” for at least 6 months. Women were considered menopausal hormone therapy (MHT) users if they used drugs included in ATC classification G03C “Estrogens” or G03F “Progestogens and estrogens in combination” for at least 6 months. 45% of the women were MHT users, and the MHT users were older on average than nonusers. Median age of MHT users at end of follow up was 70.6 years and median age of nonusers at end of follow up was 60.1 years.

### Covariates

Covariates included body mass index (BMI, kg/m^2^) and smoking status as well as sequencing batch. Individuals were classified as smoker or nonsmoker depending on whether the individual had ever been a smoker. Smoking information was not available for 43% of the population and BMI information was missing for 30%. 51% of the population had both smoking and BMI information available. Because of the large amount of missing data, the main analyses were done without smoking and BMI as a covariates, and separate analyses were carried out for the subpopulation that had smoking and BMI information available. Table 1 compares the complete dataset to the subpopulation. The first 10 genetic principal components (PC) of ancestry were used as covariates in the analyses to adjust for any genetic stratification that may occur in the study population [30].

**Table 1.** Characteristics of women in the whole data set and in the subpopulation that had BMI and smoking information available.

|  | Complete dataset | Subpopulation |
| --- | --- | --- |
| n | 184 132 | 93 931 |
| Age in years at end of follow up, mean (stdev) | 65.5 (13.5) | 65.8 (13.4) |
| BMI in kg/m<sup>2</sup>, mean (stdev) | na | 27.4 (5.6) |
| smoker % | na | 39 % |
| MHT user % | 45 % | 44 % |
| Systemic hormonal contraceptive user % | 17 % | 17 % |

### Statistical Analyses

We calculated associations between standardized polygenic scores and selected endpoints using Cox proportional hazards models and R packages *survival* [31] and *survminer* [32]. We used age as the time scale. The individual follow-up intervals started at 40 years and continued until first recorded endpoint event, death, emigration, or end-of-follow-up (December 31, 2021). We reported the results as hazard ratios (HRs) with 95% confidence intervals (CIs).

We assessed the proportional hazards assumptions by inspecting Schoenfeld residuals. We detected a time dependence for covariates BMI and smoking for all endpoints. Also MHT use and systemic contraceptive use showed time dependence. Time dependence can introduce inferential bias if a hazard ratio is not constant over the follow-up period. To meet model assumptions, we performed additional Cox analyses that stratified BMI and smoking. For the stratified analysis, BMI was divided into quantiles.

Analyses were conducted using R (version 4.2.3) software; statistical significance was set at *P* < 0.05.

## Results

### Endogenous hormonal exposure

During MegaPRS training in UK Biobank, the selected models showed moderate predictive performance for their corresponding traits (pseudo-test correlation r=0.30 for age at menarche, r=0.26 for age at menopause, and r=0.25 for length of the reproductive period), supporting their use as genetic proxies in downstream association analyses.

Figure 1 summarizes the main analyses of the associations between PGSes and endpoints for the complete study samples. Higher age at menopause PGS and higher reproductive period PGS were weakly associated with increased risk of stroke: adjusted HR was 1.02 [1.00–1.03] for both. There were no significant associations between age at menopause PGS and reproductive span PGS with hypertension or CHD events. Higher age at menarche PGS was associated with a lower risk of hypertension (HR 0.96 [0.95–0.97]) and CHD events (HR 0.97 [0.96–0.99]) but not associated with stroke. Hazard ratios are expressed per standard deviation of the respective PGS.

**Figure 1.**
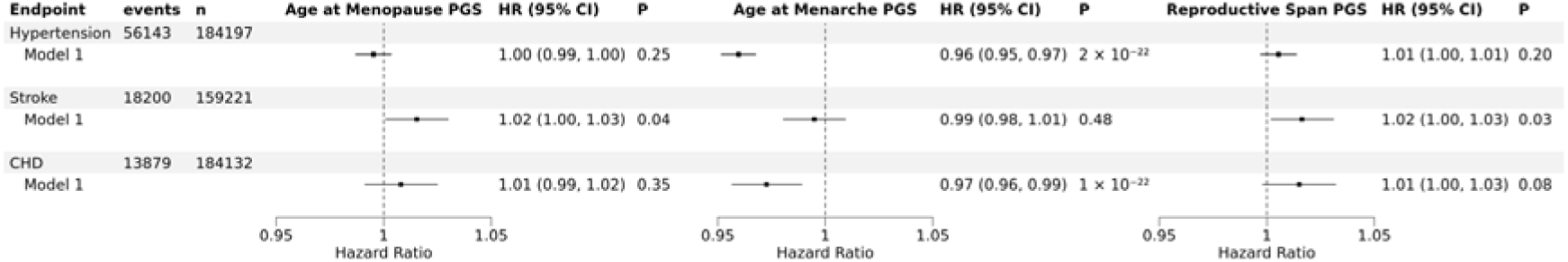
Associations of polygenic scores for age of menopause, age of menarche and length of reproductive period with hypertension, stroke, and coronary heart disease events. Multivariable Cox regression analysis: Model is adjusted for 10 genetic principal components and sequencing batch. Hazard ratios are expressed per standard deviation change of the respective PGS.

Figure 2 shows the smoking and BMI adjusted results for the subpopulation. Higher age at menopause PGS and reproductive period PGS were associated with higher stroke risk: adjusted HR was 1.03 [1.01–1.05] for both. After the smoking and BMI adjustments, age at menarche PGS was no longer associated with hypertension. Supplementary Figure 1A shows that this change resulted from adding the BMI adjustment. Supplementary Figure 1 also shows the results of stratified analyses (model 5) to be nearly identical to the results with unstratified BMI and smoking (model 4).

**Figure 2.**
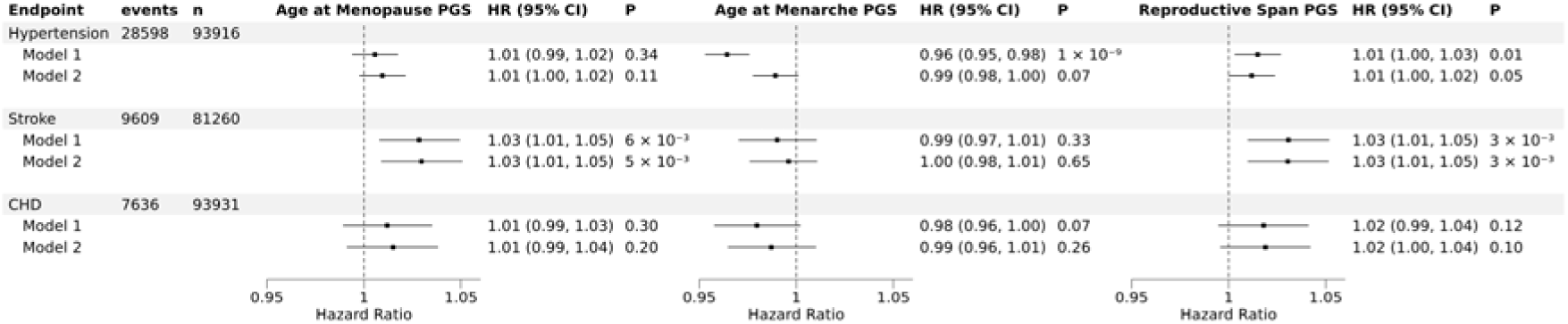
Results for the population who had BMI and smoking information available. Associations of polygenic scores for age of menopause, age of menarche and length of reproductive period with hypertension, stroke, and coronary heart disease events. Multivariable Cox regression analysis: Model 1 is adjusted sequencing batch and 10 genetic principal components. Model 2 is model 1 plus BMI and smoking. Hazard ratios are expressed per standard deviation change of the respective PGS.

### Exogenous hormonal exposure

Figure 3A shows the hazard ratios of MHT users compared to nonusers for the complete study sample and Figure 3B shows the smoking and BMI adjusted results for the subpopulation. In the whole population, MHT use was associated with lower risk of hypertension (HR 0.92 [0.91–0.94]), stroke (HR 0.82 [0.80–0.85]), and CHD events (HR 0.77 [0.74–0.80]). In the smoking and BMI adjusted results, MHT use was associated with lower risk of stroke (HR 0.85 [0.82 – 0.89]) and CHD (HR 0.80 [0.76 – 0.84]). After the BMI adjustment the association of MHT with the risk for hypertension as no longer significant.

**Figure 3.**
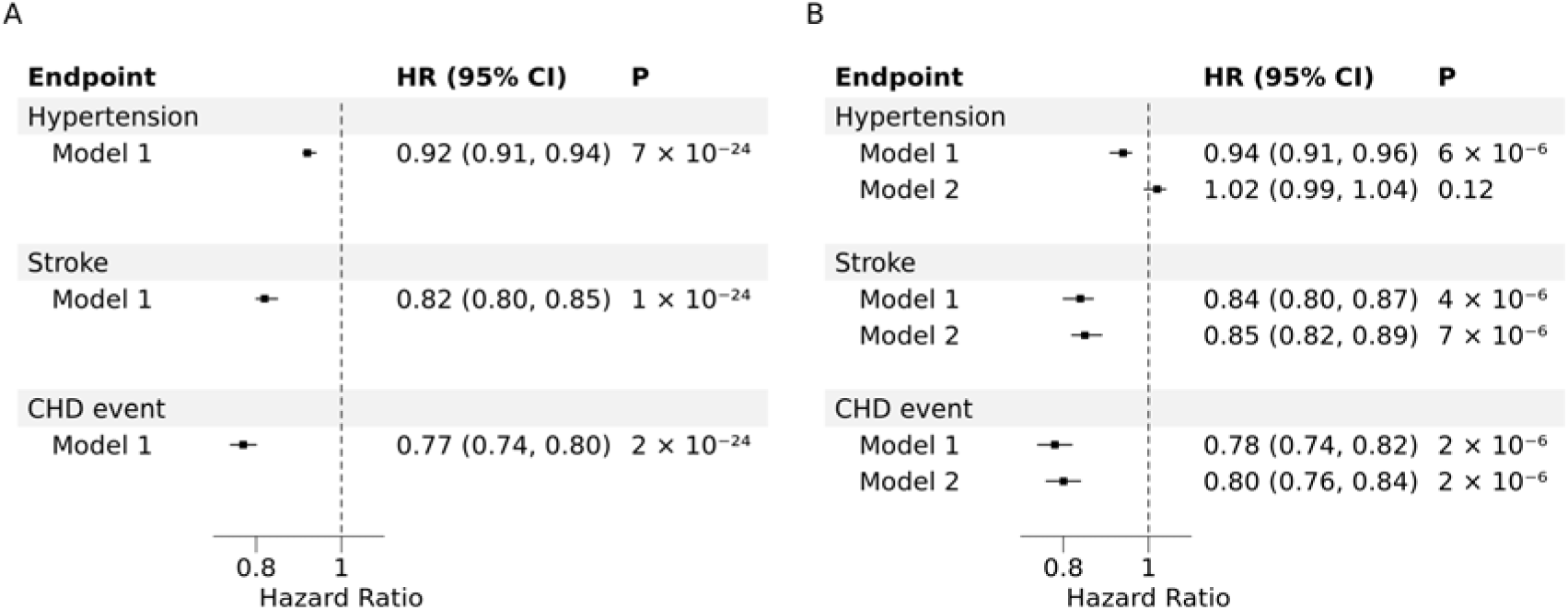
Hazard Ratios for hypertension, stroke, and CHD events of MHT users with respect to nonusers. A) Main data set analysis. Cox regression model 1 is adjusted for 10 genetic principal components and sequencing batch. B) Subpopulation analysis for the women who had BMI and smoking information available. Cox regression model 1 is adjusted for sequencing batch and 10 genetic principal components. Model 2 is model 1 plus BMI and smoking.

Figure 4A shows the hazard ratios for the complete dataset of systemic hormonal contraceptive users compared to nonusers and Figure 4B shows the smoking and BMI adjusted results for the subpopulation. In the complete dataset, systemic hormonal contraceptive use was associated with lower risk of hypertension (HR 0.90 [0.88–0.92]), stroke (HR 0.83 [0.80–0.86]), and CHD events (HR 0.82 [0.78–0.86]). Also in the adjusted results for the subpopulation, systemic hormonal contraceptive use was associated with lower risk of hypertension (HR 0.96 [0.93 – 0.99]), stroke (HR 0.84 [0.80 – 0.99]) and CHD events (HR 0.83 [0.78–0.89]). There were no significant interactions between BMI and smoking or any PGSes.

**Figure 4.**
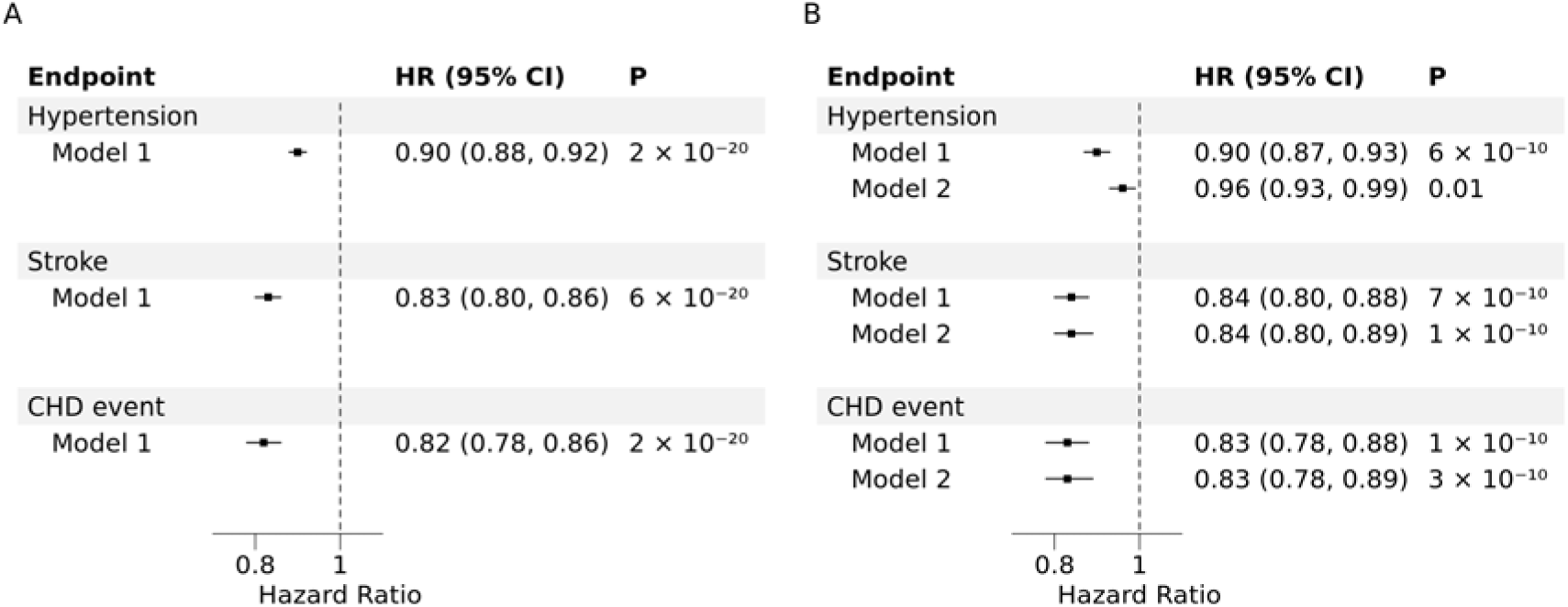
Hazard Ratios for hypertension, stroke, and CHD events of contraceptive users with respect to nonusers. A) Main data set analysis. Cox regression model 1 is adjusted for 10 genetic principal components and sequencing batch. B) Subpopulation analysis for the women who had BMI and smoking information available. Cox regression model 1 is adjusted for sequencing batch and 10 genetic principal components. Model 2 is model 1 plus BMI and smoking.

## Discussion

We found only weak associations between PGSes representing lifetime endogenous estrogen exposure and CVDs, while exogenous estrogens appeared to be beneficial for cardiovascular health in a large sample of Finnish women. According to our main analyses, an increase in age at menarche PGS was weakly associated with lower risk for hypertension and CHD events, which is consistent with prior studies associating early menarche with higher risk of CVDs [33]. While our initial results showed age at menarche PGS to be associated with CHD events, in the subpopulation analysis, the association was no longer significant. Our initial results also showed age at menarche PGS to be associated with hypertension, but after adjusting with BMI, the association was no longer significant. Since earlier menarche is associated with higher BMI [34], our results are consistent with higher BMI increasing the risk of hypertension in individuals with earlier menarche.

In our study, higher age at menopause PGS and higher reproductive period PGS were associated with a higher number of stroke events. Both scores were coded such that higher values reflect a genetically predicted later age at menopause or a longer reproductive period. Therefore, our findings suggest that women with longer lifetime endogenous estrogen exposure are at a higher risk to experience strokes. This runs counter to large observational analyses in which shorter reproductive span has generally been associated with higher cardiovascular risk – for example a pooled analysis of 12 cohorts [33] reported women with a reproductive life span of less than 30 years to have a 75% higher risk of stroke than those with a reproductive life span of 36-38 years. In another pooled analysis [15], however, there was no significant association between age of menopause and stroke. It should be taken into account that age at menopause and reproductive span are complex traits with substantial genetic and non-genetic contributions. The counterintuitive direction of our results and mixed earlier observations are plausibly explained by the fact that these polygenic scores capture genetic pathways that only partially overlap with true lifetime female hormone exposure. In other words, age at menopause and reproductive period PGSs may index biological processes (e.g., DNACdamage response and broader neuroendocrine or metabolic axes [35]) that influence vascular risk independently of, or in addition to, endogenous estrogen exposure, thereby allowing PGSCbased associations to diverge from observational findings.

About half of the variance in menopausal timing is estimated to be genetic [36], while non-genetic correlates include smoking, nutrition, adiposity and socioeconomic conditions. Smoking is the strongest non-genetic factor, and can advance menopause by 1 year [37]. Higher levels of education and socioeconomic position are associated with later menopause [37, 38]. Being overweight is associated with later menopause [37], while chronic stress is associated with earlier menopause [38]. Race and ethnicity provide a mixture of genetic and societal contributions, as age at menopause has been observed to be lowest among women from African, Latin American, Asian, and Middle Eastern countries and highest in women from Europe and Australia [37]. Our study was conducted in a Finnish population, which is relatively homogenous, thus, within our study, genetic ancestry diversity is limited. Previous observational studies evaluate the realised phenotype (the net result of genes plus environment), whereas our PGSs specifically target the genetic component. This difference matters because several nonCgenetic determinants shift age at menopause earlier and are also themselves linked with worse cardiometabolic profiles. A phenotypic association between earlier menopause and higher CVD risk can therefore, in part, reflect these environmental and social pathways that a PGS does not capture.

Most prior studies linking menopausal timing to cardiovascular outcomes focus on early (< 45 years) and premature (< 40 years) menopause. PGSes built from common variants tend to best predict variation within the normal range, while performance at the distribution tails is typically weaker. Accordingly, PGSes may underpresent the biology specific to extremely early menopause that drives much of the observational risk signal. Our results therefore suggest that although the cardiovascular vulnerability observed for women with early menopause likely reflects insufficient lifetime endogenous estrogen exposure at the phenotypic level, a genetic predisposition to a later menopause or a longer reproductive span – captured by current PGSes – does not equate to cardioprotection across the life course.

In our study, menopausal hormone therapy use was associated with a lower risk of stroke and major CHD events. After adjusting for smoking and BMI, our results showed the risk of hypertension to be increased for MHT users. It appears that BMI was a confounder in the unadjusted results. MHT users had a lower BMI than nonCusers, and because BMI is strongly associated with hypertension risk, failing to account for this imbalance may have masked the underlying association between MHT use and hypertension. The result regarding CHD was expected, as some prior studies [39, 40] have found MHT to decrease risk for CHD. Our results showing decreased stroke risk for women who had used MHT was unexpected. Several previous studies show increased stroke risk with current MHT use [24, 25, 41]. However, the current use of MHT and effects from long-term use are separate issues, and there need not be a conflict with our results and previous studies. According to one study [26], the increase in stroke risk is largest during the first year of MHT use and declines with continued use. In addition, the formulation, dose and route of delivery of MHT make a difference [24, 27]. The estradiol-based formulations that are more prevalent in Europe have been associated with lower risk of stroke than formulations with conjugated equine estrogen [42].

We found that systemic hormonal contraceptive use was associated with lower risk for hypertension, stroke and CHD events. This is somewhat contradictory to prior results that show an increased risk of stroke with oral contraceptive pill use [22, 23]. However, the risk of stroke appears to depend on estrogen dose [23] and progestin-only pills were not associated with increased stroke risk [43]. Further on, it has been suggested [26] that oral contraceptives elevate the stroke risk mainly during the first year after initiation but not afterwards, and that the risk is reversed [23, 26] after cessation of oral contraceptive use. We looked at hormone exposure over the long term, and classified women as contraceptive users if they had ever used systemic hormonal contraceptives.

Our study has some limitations. This was a retrospective study and may suffer from selection bias. The high amount of missing smoking and BMI data is another limitation. There are many potential confounders that were not considered, including the use of other medications and potential comorbid conditions. We only looked at whether or not participants had used MHT or systemic hormonal contraceptives, without considering the length of use, formulation, dose, and route of delivery.

## Conclusions

Higher age at menopause PGS and reproductive span PGS were weakly associated with elevated risk of stroke. Because stroke risk is known to increase with lower age at menopause and shorter reproductive span, the contribution from common genetic variants appears to be clinically less important. MHT use was associated with decreases in stroke and CHD event risks. Systemic hormonal contraceptive use was associated with a lower risk of hypertension, stroke, and CHD over the course of the lifespan. Given that previous literature shows increased stroke risk with current use of exogenous estrogen, our results showing decreased long-term stroke risk with MHT use and systemic hormonal contraceptive use require further validation from future studies.

## Supporting information

FinnGen authors

## Data Availability

Data are available upon reasonable request. Researchers can apply to use the FinnGen resource and access the data used.

## Nonstandard Abbreviations and Acronyms

BMI: body mass index
CVD: cardiovascular disease
CHD: coronary heart disease
GWAS: genome-wide association study
HR: hazard ratio
MHT: menopausal hormone therapy
PC: principal component
PGS: polygenic score
UKBB: UK Biobank

## Acknowledgments

We want to acknowledge the participants and investigators of the FinnGen study. The FinnGen project is funded by two grants from Business Finland (HUS 4685/31/2016 and UH 4386/31/2016) and the following industry partners: AbbVie Inc., Alnylam Pharmaceuticals, Inc., AstraZeneca UK Ltd, Bayer AG, Biogen MA Inc., Boehringer Ingelheim International GmbH, Bristol Myers Squibb Inc. (and Celgene Corporation & Celgene International II Sàrl), Genentech Inc., GlaxoSmithKline Intellectual Property Development Ltd., Johnson&Johnson Innovative Medicine Inc., Maze Therapeutics Inc., Merck Sharp & Dohme LCC, Novartis AG, Pfizer Inc. and Sanofi US Services Inc. Following biobanks are acknowledged for delivering biobank samples to FinnGen: Auria Biobank (www.auria.fi/biopankki), THL Biobank (www.thl.fi/biobank), Helsinki Biobank (www.helsinginbiopankki.fi), Biobank Borealis of Northern Finland (https://www.ppshp.fi/Tutkimus-ja-opetus/Biopankki/Pages/Biobank-Borealis-briefly-in-English.aspx), Finnish Clinical Biobank Tampere (www.tays.fi/en-US/Research_and_development/Finnish_Clinical_Biobank_Tampere), Biobank of Eastern Finland (www.ita-suomenbiopankki.fi/en), Central Finland Biobank (www.ksshp.fi/fi-FI/Potilaalle/Biopankki), Finnish Red Cross Blood Service Biobank (www.veripalvelu.fi/verenluovutus/biopankkitoiminta), Terveystalo Biobank (www.terveystalo.com/fi/Yritystietoa/Terveystalo-Biopankki/Biopankki/) and Arctic Biobank (https://www.oulu.fi/en/university/faculties-and-units/faculty-medicine/northern-finland-birth-cohorts-and-arctic-biobank). All Finnish Biobanks are members of BBMRI.fi infrastructure (https://www.bbmri-eric.eu/national-nodes/finland/). Finnish Biobank Cooperative -FINBB (https://finbb.fi/) is the coordinator of BBMRI-ERIC operations in Finland. The Finnish biobank data can be accessed through the Fingenious^®^ services (https://site.fingenious.fi/en/) managed by FINBB.

---

**Supplementary Figure 1.**
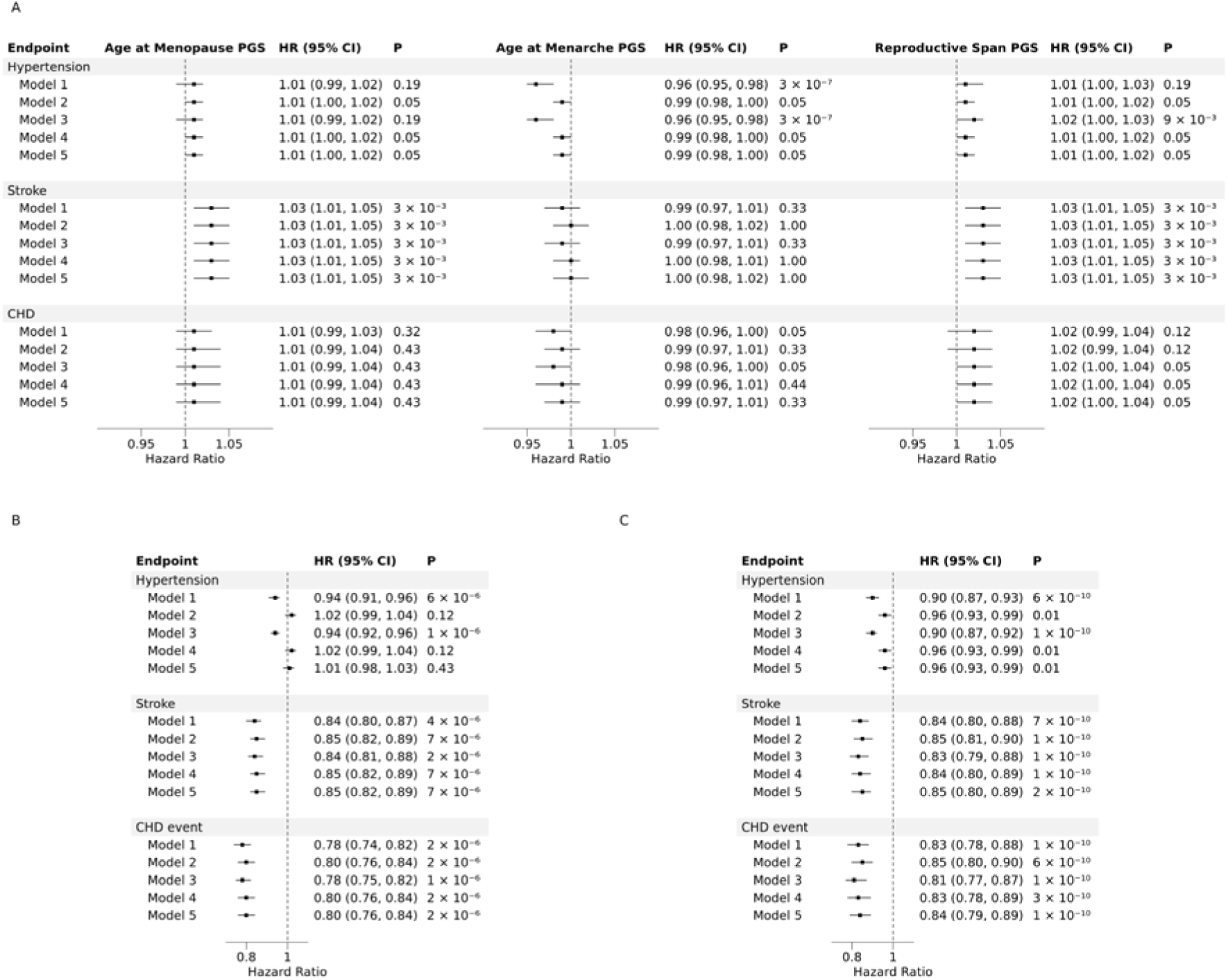
Supplementary results for the population who had BMI and smoking information available. A) Associations of polygenic scores for age of menopause, age of menarche and length of reproductive period with hypertension, stroke, and coronary heart disease events. Hazard ratios are computed for each standard deviation for the respective PGS. B) results for MHT users with respect to nonusers and C) results of systemic contraceptive users with respect to nonusers. Multivariable Cox regression analysis: Model 1 is adjusted for sequencing batch and 10 genetic principal components. Model 2 is model 1 plus BMI. Model 3 is model 1 plus smoking. Model 4 is model 1 plus BMI and smoking. Model 5 is model 1 plus stratified BMI and smoking.

**Supplementary Table 1.** ICD codes included in Finngen endpoint definitions I9_HYPTENS, C_STROKE and I9_CHD.

| Endpoint | Hospital discharge register codes |  |  |  | Cause of death register codes |  |  |  |
| --- | --- | --- | --- | --- | --- | --- | --- | --- |
|  | ICD_10 | ICD_9 | ICD_8 | Excluded | ICD_10 | ICD_9 | ICD_8 | Excluded |
| I9_HYPTENS | I10, I11, I12, I13, I15, I674 | 4019X, 4029A, 4029B, 4039A, 4040A, 4059A, 4059B, 4372A, 4059X | 400, 401, 402, 403, 404 |  | I10, I11, I12, I13, I15, I674 | 4019X, 4029A, 4029B, 4039A, 4040A, 4059A, 4059B, 4372A, 4059X | 400, 401, 402, 403, 404 |  |
| C_STROKE | I60, I61, I63, I64, G45 | 430, 431, 4330A, 4331A, 4339A, 4340A, 4341A, 4349A, 435, 436 | 430, 431, 433, 434, 435, 436 | ICD_10 G454, ICD_10 I636, ICD_8 43101, ICD_8 43191 |  | 430, 431, 4330A, 4331A, 4339A, 4340A, 4341A, 4349A, 435, 436 | 430, 431, 433, 434, 435, 436 | ICD_10 G454, ICD_10 I636, ICD_8 43101, ICD_8 43191 |
| I9_CHD | I200, I21, I22 | 410, 4110 | 410, 4110 |  | I2[1-5], I46, R96, R98 | 41[0-4], 798 | 41[0-4], 798 | ICD_9 7980A |

